# Telmisartan for treatment of Covid-19 patients: an open randomized clinical trial - A preliminary report

**DOI:** 10.1101/2020.08.04.20167205

**Authors:** Mariano Duarte, Facundo Pelorosso, Liliana Nicolosi, M. Victoria Salgado, Héctor Vetulli, Analía Aquieri, Francisco Azzato, Mauro Basconcel, Marcela Castro, Javier Coyle, Ignacio Davolos, Eduardo Esparza, Ignacio Fernandez Criado, Rosana Gregori, Pedro Mastrodonato, María C. Rubio, Sergio Sarquis, Fernando Wahlmann, Rodolfo P. Rothlin

## Abstract

**Background:** Covid-19 is associated with respiratory-related morbidity and mortality. Angiotensin receptor blockers (ARB) have been postulated as tentative pharmacological agents to treat Covid-19-induced inflammation.

**Methods:** This is a randomized, two-arm, open, multicenter trial. Participants were 18 years or older and had been hospitalized with confirmed Covid-19 with 4 or fewer days since symptom onset. Exclusion criteria included intensive care unit admission prior to randomization and ARB or angiotensin converting enzyme inhibitors use. Treatment arm received telmisartan 80 mg bid during 14 days plus standard care; control arm received standard care. Primary outcome were differences in C-reactive protein levels at days 5 and 8. Secondary outcomes included time to discharge evaluated at 15 days and death at 30 days post randomization.

**Results:** This interim analysis included 40 patients in telmisartan and 38 in control groups. CRP levels in the control and telmisartan groups were 51.1±44.8 mg/L vs 24.2±31.4 mg/L at day 5 (mean ± SD; n=28 and n=32, p<0.05), and 41.6±47.6 mg/L vs 9.0±10.0 mg/L at day 8 (mean ± SD; n=16 and n=13; p<0.05), respectively. Telmisartan treated patients had statistically significant lower time to discharge (log-rank test p=0.0124, median time: 15 days in control group vs 9 days in telmisartan group). Mortality at day 30 was 11.76% in control group vs 5.26% in telmisartan group (p=0.41).

**Conclusions:** In this study, ARB telmisartan, a well-known inexpensive safe antihypertensive drug, administered in high doses, was superior to standard care demonstrating anti-inflammatory effects and improved morbidity in hospitalized patients infected with SARS-CoV-2 (<u>NCT04355936</u>).

## Introduction

The SARS-CoV-2 virus enters the airway and binds the host cell (alveolar type 2) through the interaction of the structural protein S (Spike) with the protein membrane ACE2 (angiotensin-converting enzyme 2)^1^. The virus-ACE2 complex is internalized by endocytosis effectively sequestering (apparent down-regulation) ACE2 which in turn loses its function catalyzing the degradation of angiotensin ll to angiotensin 1-7. Angiotensin II acting on AT1 receptors causes vasoconstriction, apoptosis, proinflammatory effects, and fibrosis. Angiotensin 1-7 acting on Mas receptors causes opposite effects: it mediates vasodilation and anti-inflammatory actions ^2^. Coronavirus disease 2019 (Covid-19), the disease caused by SARS-CoV-2, is associated with significant respiratory-related morbidity and mortality, as well as elevation of systemic inflammatory biochemical markers ^3^. Among them, one of the most relevant is C-reactive protein (CRP) whose serum levels can be used as an independent factor to predict the disease severity and progression ^4,5^. CRP is a pentameric protein induced by IL-6 in the liver and the expression level increases rapidly and significantly during acute inflammatory responses^6^. In Covid-19 patients serum levels of CRP rise during the initial phase of infection, with continued increments during the first week of the disease to then fall dramatically in a few days to normal values if recovery ensues ^7,8^. CRP has been shown to correlate with clinical course and in critically ill patients can even rise or remain high ^4^.

It has been proposed that in Covid-19 patients the loss of ACE2 function in alveolar cells results in a deviation of the homeostatic balance of the renin angiotensin system leading to increased tissue concentration of angiotensin II and reduced levels of its physiological antagonist angiotensin 1-7^3^. Angiotensin II can promote apoptosis in alveolar cells, which, in turn, initiates an inflammatory process with release of proinflammatory cytokines, establishing a self-powered cascade ^9^ leading, in severe cases, to Acute Respiratory Distress Syndrome (ARDS) ^10^. Elevation of angiotensin II in other tissues seems to play a role in promoting inflammation and tissue injury (myocarditis, renal injury, etc). Angiotensin receptor blockers (ARBs), a well-known anti-hypertensive drug group that blocks AT1 receptor, have been postulated as tentative pharmacological agents to treat Covid-19-induced lung inflammation ^11^. Data from retrospective studies from Covid-19 patients have provided some evidence to support that hypothesis ^12^. However, no conclusive data from a prospective randomized trial on the use of ARBs on Covid-19 patients are available. Pharmacological analysis made by Rothlin et al. ^13^ has postulated that telmisartan may be the best candidate to study. Therefore, the objective of this study was to assess whether telmisartan 80 mg b.i.d. would be effective in reducing lung inflammation and CRP levels at 5 and 8 days of treatment in Covid-19 hospitalized patients. We present here a preliminary report of that study.

## Methods

### Trial design and settings

We conducted this two-arm, multicenter, randomized, open-label, controlled trial at two academic hospitals in Ciudad de Buenos Aires, Argentina: Hospital de Clínicas “José de San Martín” or site 1 (HCJSM, University of Buenos Aires main hospital) and Hospital Español de Buenos Aires or site 2 (HEBA, a community hospital). The protocol was approved by the ethics committee at HCJSM and the institutional review board at HEBA. The trial was funded by the participating hospitals. Laboratorio Elea Phoenix S.A. donated and supplied the trial drug and provided administrative support for registration of this trial at www.ClinicalTrials.gov. Laboratorio Elea Phoenix S.A. had no role in the conduct of the trial, the analysis, or the decision to submit the manuscript for publication.

### Participants

The trial included participants who were 18 years of age or older and who had been hospitalized with PCR-confirmed Covid-19 infection with 4 or fewer days elapsed since symptom onset. Exclusion criteria were: admission to Intensive Care Unit (ICU) prior to randomization, illness symptoms beginning more than 4 days prior to randomization, pregnancy, breast feeding, major hypersensibility to ARBs, systolic blood pressure < 100mmHg, serum potassium greater than 5.5 mEq/L, AST and/or ALT > 3 times the upper limit of normal, serum creatinine higher than 3 mg/dL, treatment with angiotensin converting enzyme inhibitor (ACEi) or ARB at admission. All the patients provided written informed consent before randomization.

### Randomization and intervention

Patients were randomly assigned in a 1:1 ratio to receive standard care (control group) or standard care plus telmisartan 80 mg twice daily for 14 days. Simple randomization was performed using the GraphPad QuickCalcs Web site by a statistician with no contact with patient care. Patients who received plasma from convalescent patients were censored from the date of plasma administration onwards. Guidance was provided to the investigators about how to adjust or interrupt treatment according to side effects and laboratory abnormalities.

### Outcomes

Reductions of C reactive protein levels at day 5 and 8 after randomization were chosen as primary outcome. Secondary outcomes included admission to ICU within 15 and 30 days from randomization, occurrence of mechanical ventilation within 15 and 30 days from randomization, death within 15 and 30 days from randomization, composite occurrence of admission to ICU, mechanical ventilation or death within 15 and 30 days from randomization, proportion of patients requiring supplemental oxygen at day 15 (or with an oxygen saturation lower than 96% while breathing room air), time from randomization to discharge up to day 15 from randomization, and significative differences in serum lactate dehydrogenase levels at day 5 and 8. All the trial outcomes were assessed by the site investigators, who were aware of the trial-group assignments.

### Sample size calculation and protocol changes

For sample size calculations, we used our main outcome level as reference (CRP), and a repeated measures model. Calculations were done using the GLIMMPSE (General Linear Mixed Model Power and Sample Size) software ^14^, freely available at https://glimmpse.samplesizeshop.org/. We determined a 0.80 power and a type I error rate of 0.05, and chose the Hotelling Lawley Trace test.

We assumed an initial CRP level of 60 mg/L in both groups, with an elevation on day 5 in the control group (up to 72 mg/L, 20% more) and a descend in the telmisartan group to 36 mg/L (40% less). We then assumed that the mean value decreased at day 8 in both groups. Accounting for variability on these assumptions, we used a scale factor of 0.5 for the mean and 2 for the standard deviation. Further detail is provided in the Statistical analysis plan in the trial protocol.

We obtained a total population sample of 390 participants (195 in each group), which we roughly approximated to 400 (200 in each group).

Initial design included CRP level comparison at day 8 and 15. Reports made after the writing of the original version of the protocol as well as observations made by the research team showed that CRP levels may fall rapidly in recovering patients^5,8^. Therefore, original endpoint measuring CRP levels at day 15 after randomization was replaced by an earlier timepoint at day 5 to better capture the dynamic of CRP changes and its potential modification by telmisartan treatment. Additional support of this change was brought by the length of stay of patients which was often shorter than 15 days, leading to an excessive amount of missing data. Although changes in the primary outcome are not common for diseases that are well understood, it is recognized that in some trials, such as those involving poorly understood diseases, circumstances may require a change in the way an outcome is assessed or may necessitate a different outcome^15^.

Composite occurrence of admission to ICU, mechanical ventilation or death between randomization and 15 and 30 days, proportion of patients requiring supplemental oxygen at day 15, time to discharge from randomization at 15 days were also added as secondary outcomes at that point.

### Statistical analysis

This is a two arm, open label, randomized trial testing a superiority hypothesis with a two-sided type I error rate of 0.05 (refer to the trial protocol for more information on statistical analysis plan).

Descriptive analysis was performed using the appropriate summary statistics (e.g., proportions for categorical data, means with 95% confidence intervals for continuous data, median for time-to-event data). Comparison of C-reactive protein and LDH levels at day 1, 5 and 8 were analyzed by fitting a mixed model. Mixed-effects model analysis for repeated measures was followed by a Holm-Šídák multiple comparison test. Analysis of time to discharge was done calculating proportions using the Kaplan-Meier method, and the resulting curves were compared by a log-rank test. Differences in proportions (ICU, mechanical ventilation, death, need for oxygen supplementation at day 15) were compared by Chi-square test. Analyses were performed using GraphPad Prism version 8.4.3 (686) for Windows. We planned one interim analysis to be conducted on July 31^st^ 2020.

Additional information can be found at the Supplementary Appendix.

## Results

### Characteristics of the participants

We recruited 82 participants with confirmed Covid-19. The numbers of enrolled patients were 66 and 16 at site 1 and site 2, respectively. A total of 41 patients were randomly assigned to receive telmisartan and 41 patients to receive standard care (control group) (Figure 1). The first patient underwent randomization on May 14, 2020; the last patient before interim analysis underwent randomization on July 30, 2020. Four patients were excluded after randomization (3 patients in the control group met exclusion criteria and 1 patient did not receive telmisartan treatment). Four patients in the control group and two in the treatment group were censored before day 14 because administration of plasma from convalescent patients.

**Figure 1.**
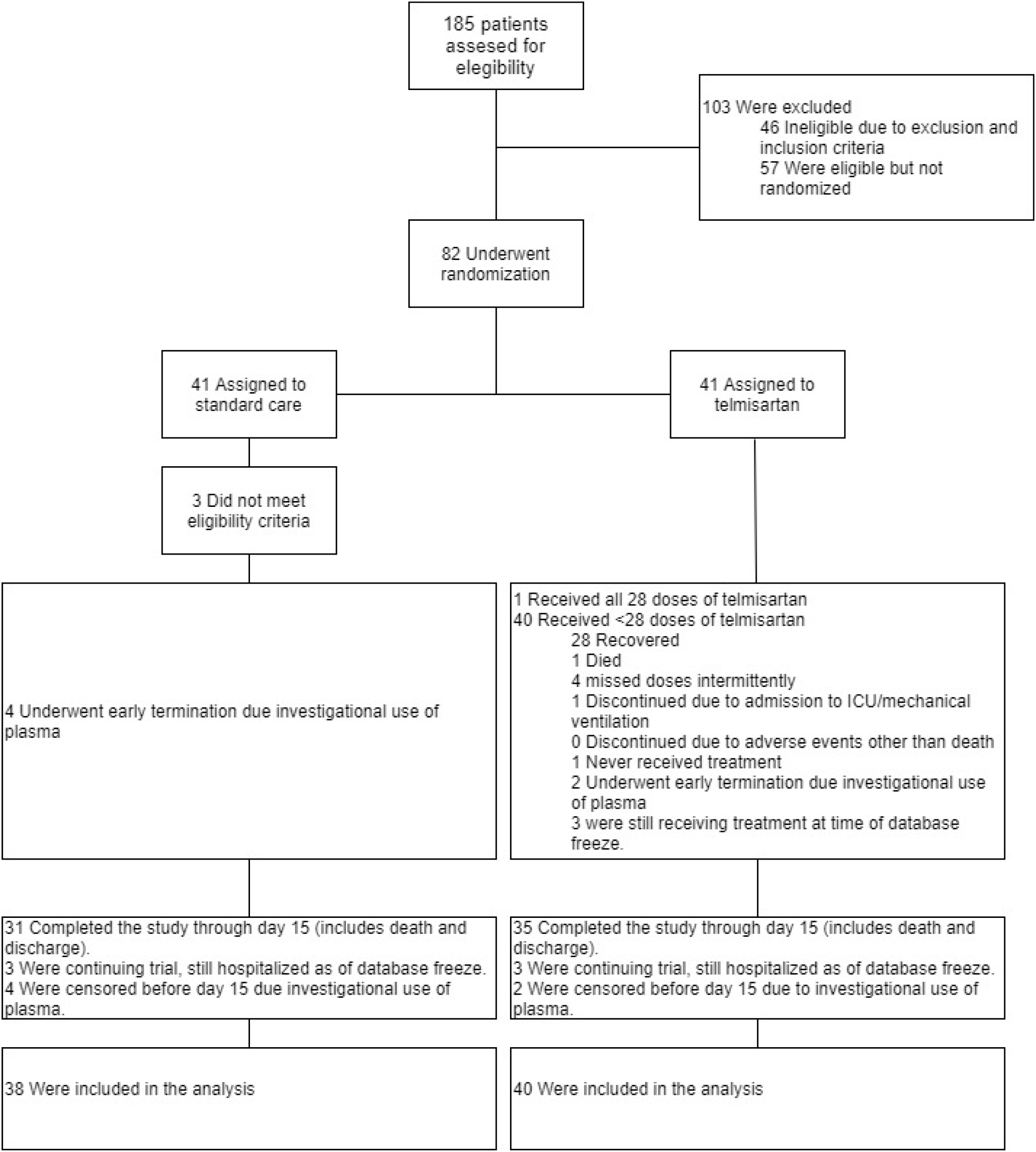
Enrollment and randomization.

At the time of the database freeze (July 31^st^, 2020), a total of 3 patients in the control group and 3 patients in the telmisartan group had not reached day 15 after randomization.

No statistically significant imbalances were observed between groups at baseline. Demographic and clinical characteristics of participants are depicted in Table 1.

**Table 1.** Demographic and clinical characteristics of participants at baseline, by treatment group.

|  |  |  |
| --- | --- | --- |
| **Characteristics** | **control (n=38)** | **telmisartan (n=40)** |
| Age -years | 63.8 ± 18.7 (38) | 60.1 ± 17.8 (40) |
| Female - no (%) | 17 (44.7) | 13 (32.5) |
| Coexisting conditions - no (%) |  |  |
| Hypertension | 10 (26.3) | 14 (35.0) |
| COPD | 2 (5.3) | 7 (17.5) |
| Diabetes | 5(13.2) | 4 (10.0) |
| Obesity | 2 (5.3) | 8 (20.0) |
| Dyslipemia | 4 (10.5) | 7 (17.5) |
| Stroke | 1 (2.6) | 5 (12.5) |
| Asthma | 0 (0) | 1 (2.5) |
| Chronic kidney disease | 0 (0) | 2 (5.0) |
| Clinical characteristics at admission |  |  |
| Required supplementary O<sub>2</sub> - no (%) | 30 (79.0) | 35 (87.5) |
| Respiratory rate (bpm) | 19.8 ± 3.1 (37) | 19.5± 2 (39) |
| CRP (mg/L) | 53.7 ± 56.7 (38) | 74.5 ± 72.5 (37) |
| Lymphocyte count (10<sup>3</sup>/μL) | 1.25 ± 0.1 (37) | 1.33 ± 0.1 (36) |
| Platelet count (10<sup>3</sup>/μL) | 200.8 ± 9.7 (38) | 235.3 ± 15.8 (36) |
| Neutrophil to lymphocyte ratio | 4.8 ± 0.8 (37) | 7.0 ± 1.5 (37) |
| LDH (UI/L) | 520.6 ± 267.11 (35) | 506.7 ± 30.2 (33) |
| D-Dimer (μg/mL) | 1.30 ± 0.89 (21) | 0.98 ± 0.78 (22) |
| Ferritin (ng/mL) | 1129.8 ± 290.3 (17) | 804.6 ± 132.9 (19) |
| COPD: Chronic Obstructive Pulmonary Disease; CRP: C reactive protein; LDH: Lactate Dehydrogenase |  |  |
| Plus-minus values are means ±SD; () indicates the n. All differences were not statistically significant. |  |  |

### Primary outcomes

Patients in the telmisartan group had a lower CRP serum level than patients in the control group at day 5 (control 51.1 ± 44.8 mg/L, n=28, as compared with telmisartan 24.2 ± 31.4 mg/L, n=32, p<0.05, all values are expressed as mean ± SD). Also, CRP serum levels were lower at day 8 in patients treated with telmisartan than those in the control group (control: 41.6 ± 47.6 mg/L, n=16, as compared with telmisartan: 9.0 ± 10.0 mg/L, n=13, p < 0.05) (Figure 2).

**Figure 2.**
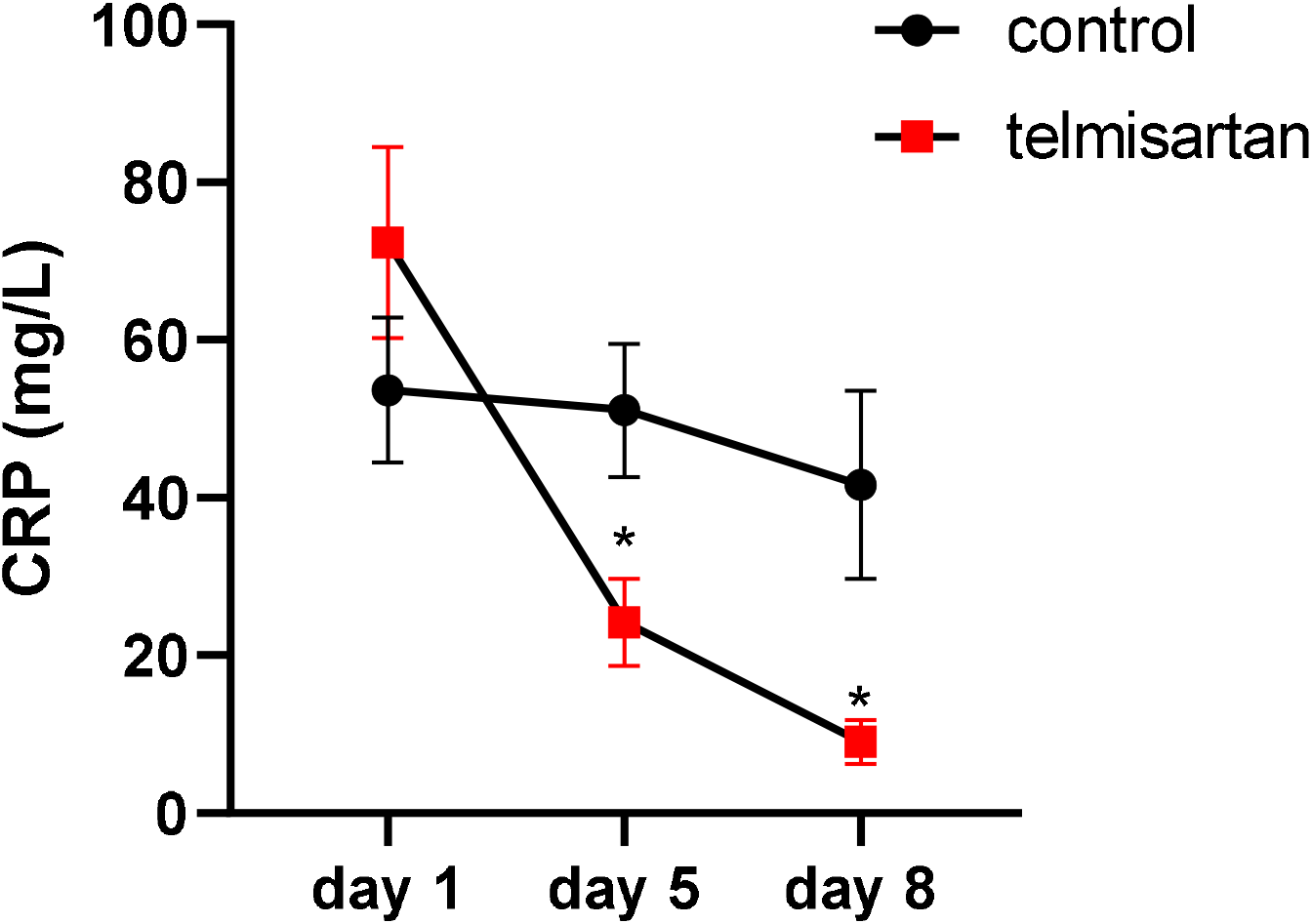
Serum CRP levels at days 1, 5 and 8 after randomization. Points represent mean ± SEM. * indicates p<0.05.

### Secondary outcomes

Results from clinical secondary outcomes are shown in Table 2. Preliminary results from the 78 patients analyzed (40 assigned to telmisartan and 38 assigned to standard care) with data available up to 15 days after randomization indicated that those who received telmisartan had a median discharge time of 9 days, as compared with 15 days in those who received standard care (Log-rank (Mantel-Cox) p=0.0124), hazard ratio (log-rank) for discharge telmisartan/control 2.02 (95% CI, 1.14 to 3.59) (Table 2 and Figure 3).

**Table 2:** Clinical Evolution at day 15, by treatment group.

| Table 2: Clinical Evolution at day 15, by treatment group |  |  |
| --- | --- | --- |
| Outcome | control | telmisartan |
| Hospital discharges by day 15 – no. (%) | 18 (56.4) | 29 (79.5) |
| Median time to discharge (days) | 15 | 9 |
| Risk of discharge telmisartan/control – HR (95% CI) | 2.02 (1.14 to 3.59) |  |
| Hospitalized patients needing supplementary O<sub>2</sub> at day 15 – no. (%) † | 13/14 (0.93) | 2/4 (0.5) * |
| *p<0.05. † or with an oxygen saturation lower than 96% while breathing room air. |  |  |

**Figure 3.**
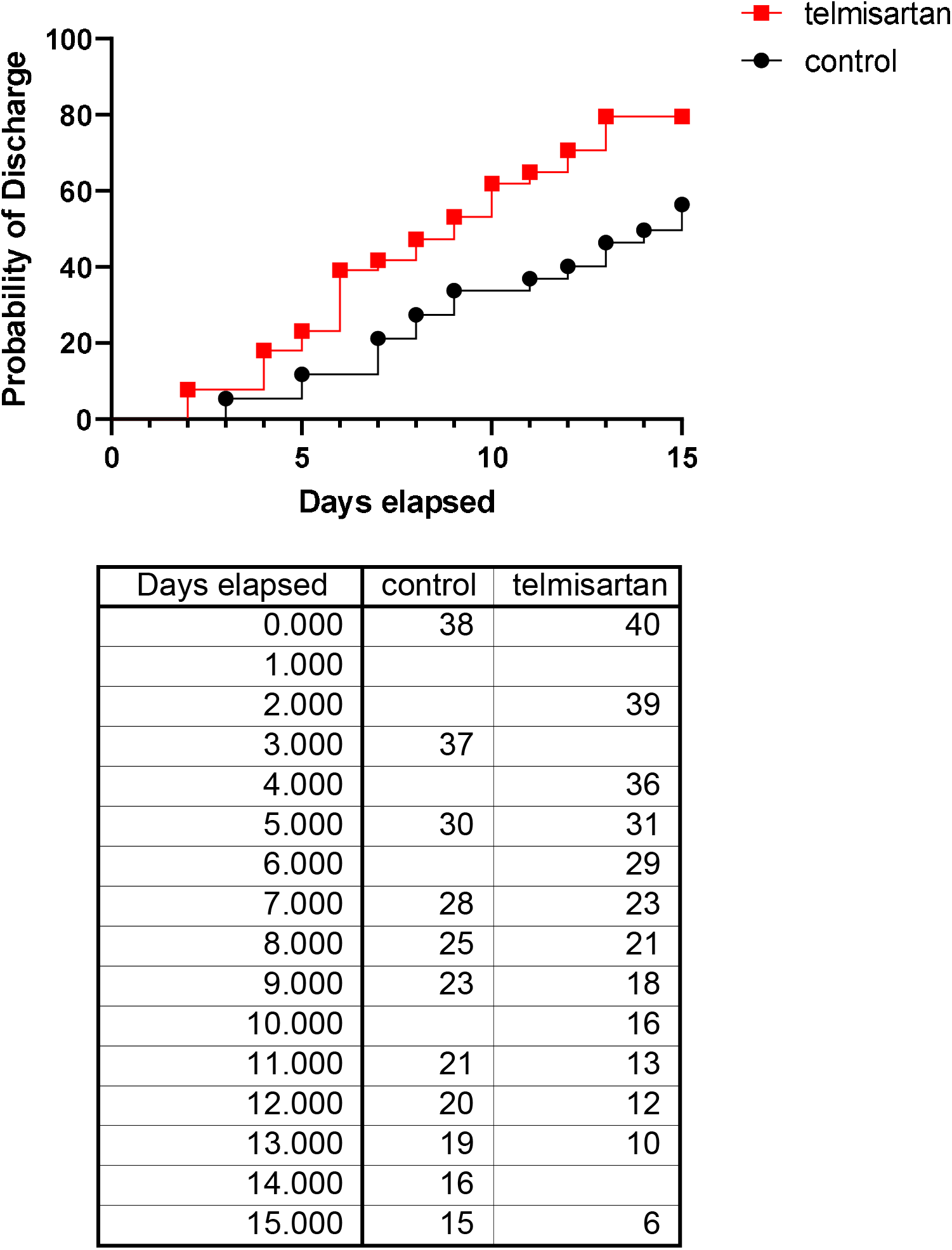
Kaplan-Meier Estimates of Time from Randomization to Discharge at day 15.

By day 30, there were 4 deaths out of 34 patients in the control group vs 2 out of 38 in the telmisartan group (11.76 % vs 5.26 %, respectively, p=0.41). No differences were observed for ICU admission, mechanical ventilation, death or the composite outcome of first occurrence of ICU admission, mechanical ventilation or death at day 15 or day 30 (Table S2). A detail of patient’s characteristics and causes of death can be found in the Supplementary Appendix (Table S3).

The proportion of inpatients needing supplementary O_2_ at day 15 was significantly higher in the control group (13 out of 14) than in telmisartan treated patients (2 out of 4; p<0.05, Table 2). Reductions observed in LDH levels at days 5 and 8 between control and telmisartan group did not reach statistical significance (day 5: control group: 511.0 ± 196.7, n=35, telmisartan group: 452.7 ± 159.6, n=28; day 8: control group: 501.8 ± 209.5, n=15, telmisartan group: 393.4 ± 60.7, n=12, p>0.05, all values are expressed as mean ± SD, Figure S1a).

No adverse events related to telmisartan were reported. No differences were observed in blood pressure between telmisartan and control group at day 5 or 8 (Table S4).

Hematological indices and additional biomarkers at day 5 and day 8 are shown in the Supplemental Appendix (Figure S1 and Table S5).

## Discussion

In the present preliminary report, despite the small number of patients studied, telmisartan decreased plasma CRP levels rapidly and in a sustained manner. At day 5, CRP levels were significantly reduced from 51.1 mg/L in the control group to 24.2 mg/L in the telmisartan treated group. Levels at day 8 remained significantly lower in the treated group (9.0 mg/L) compared to control group (41.6 mg/L). Moreover, telmisartan treatment produced an improvement in the clinical evolution of patients hospitalized with Covid-19 as evidenced by a shorter time to discharge measured during the first 15 days of hospitalization (log-rank test p=0.0124, median time to discharge: 15 days in control group vs 9 days telmisartan group). In the same line, patients still hospitalized at day 15 were less likely to require oxygen supplementation in the treated group than in the control group.

The hypothesis of the involvement of the renin angiotensin system in the inflammatory process triggered by the entry of SARS-COV-2 into the tissues (lung in first place) considers that the down regulation of ACE2 causes an imbalance which results in an elevation of angiotensin ll and a reduction in angiotensin 1-7 extracellular concentrations. Taking into account that angiotensin II has pro-inflammatory properties and that angiotensin 1-7 has anti-inflammatory properties^16^, this imbalance could induce the development of inflammatory processes AT1 receptor dependent^11,13^.

In Covid-19 patients, Liu F et al. (2020) demonstrated that plasma angiotensin II levels were linearly associated with viral load and lung injury^4^. In line with these results, also in Covid-19 patients, Villard et al (2020) show that plasma levels of aldosterone and CRP at admission were significantly higher in patients with a severe clinical course as compared to those with a mild or moderate clinical course^17^. Taking into account that aldosterone is synthesized in the adrenal cortex in response to angiotensin II, these results strongly suggest that they correspond to an increase in plasma levels of angiotensin II.

In the lung, the RAS imbalance could induce angiotensin II to trigger an inflammatory process through the stimulation of AT1 receptors in a variety of cells (pneumonocytes, macrophages, epithelial cells, and endothelial cells) releasing pro-inflammatory cytokines^16^. Among these, IL6 is particularly relevant since its plasma levels are directly related to the severity of Covid-19^4,18^ and also induces the CRP gene promoting an increase in its production. In this context, it is important to indicate that CRP serum levels are considered an independent predictive biomarker of the severity of the Covid-19 disease on admission^4^.In addition, in recent years, solid experimental information indicates that CRP has inflammatory amplifying properties in inflamed tissues ^6,14^.

To evaluate the involvement of RAS in systemic inflammation and clinical evolution of hospitalized Covid-19 patients, this protocol was designed using telmisartan, an AT1 receptor blocker. Telmisartan was chosen for its pharmacokinetic and pharmacodynamic properties ^13^. Telmisartan, which is well absorbed after oral administration, is the ARB with the longest plasma half-life (24 h), it reaches the highest tissue concentrations due to its high lipid solubility and high volume of distribution (500 L), and dissociates more slowly after binding to the AT1 receptor, causing an apparently irreversible block ^19,20^. In addition, the use of a daily dose of 160 mg was defined, higher than the maximum recommended antihypertensive, taking into account its “placebo like” adverse effect profile ^21^).

In the present study, treatment with telmisartan in patients with Covid-19 produced a significant reduction of serum CRP levels 5 and 8 days after admission (Figure 2). The differences observed between control and telmisartan group (51.1 mg/L vs 24.2 mg/L at day 5 and 41.6 mg/L vs 9.0 mg/L at day 8) might be clinically relevant considering that patients with CRP levels higher than 40 mg/L are more likely to have severe complications. Simultaneously, telmisartan was superior to standard care in significatively shortening the time to discharge in adults hospitalized with Covid-19 (Figure 3). These data, when taken together, indicate that telmisartan reduced morbidity in Covid-19 patients in this study. It has been shown that most patients only develop antibodies against SARS-COV2 after 7 days from disease onset and that, by day 15, IgM and IgG antibodies are detected in approximately 90% and 80% of the patients ^22^. Also, Sun et al. ^23^ showed that, as Covid-19 progressed, the increase of IgG specific to S protein positively correlated with a decrease of CRP in non-ICU patients. Therefore, the rapid anti-inflammatory effect of telmisartan may be highly valuable in improving the final outcome of the clinical course of Covid-19 patients.

Main limitations of the study are the lack of blinding, the exclusion of ICU patients on randomization, the low number of enrolled patients given the preliminary nature of this report, and the restriction to patients with a relatively short time from symptom onset to randomization. Therefore, additional rigorous studies are warranted to determine its value as a therapeutic tool in the Covid-19 clinical spectrum.

In synthesis, the present results constitute strong evidence in favor of the involvement of the RAS in the inflammatory process observed in hospitalized Covid-19 patients and that the ARB telmisartan, a well-known inexpensive safe antihypertensive drug, administered in high doses, demonstrates anti-inflammatory effects and improved morbidity in hospitalized patients infected with SARS-CoV-2, providing support for its use in this serious pandemia.

## Data Availability

Data of individual participants that underlie the results reported in this article, after deidentification (text, tables, figures, and appendices) will be made available upon publication for 5 years at a third-party website (DOI: 10.5281/zenodo.3970223)

http://10.5281/zenodo.3970223

## Acknowledgements

The authors would like to acknowledge Dr. Carlos R. Rojo, MD. for sharing our hypothesis and promoting institutional support for the conduction of this study.

The authors would also like to acknowledge Dr. Raúl Mejía for the critical reading of this manuscript.

## Role of study sponsors

The School of Medicine, University of Buenos Aires, provided material support through permission to use Hospital de Clínicas facilities to carry out the trial. Also, all biochemical determinations at Hospital de Clínicas were carried out at its Central Laboratory Facility. Hospital Español de Buenos Aires provided material support through permission to use its facilities to carry out the trial. Also, all biochemical determinations at this site were carried out at the Central Laboratory Facility at Hospital Español. Laboratorios Elea Phoenix provided the telmisartan tablets used for the study and provided assistance in submitting the registration of this trial to www.ClinicalTrials.com.

The sponsors had no role in the design of this study neither had any role during its execution, analyses, interpretation of the data, or decision to submit results.

**Preprint of a previous version of this manuscript has been submitted to medRxiv (https://doi.org/10.1101/2020.08.04.20167205)**.

## References

1. Wan Y, Shang J, Graham R, Baric RS, Li F. Receptor Recognition by the Novel Coronavirus from Wuhan: an Analysis Based on Decade-Long Structural Studies of SARS Coronavirus. J Virol 2020;94.

2. Paz Ocaranza M, Riquelme JA, García L, et al. Counter-regulatory renin-angiotensin system in cardiovascular disease. 2020;17:116–29.

3. Liu Y, Yang Y, Zhang C, et al. Clinical and biochemical indexes from 2019-nCoV infected patients linked to viral loads and lung injury. Science China Life sciences 2020;63:364–74.

4. Liu F, Li L, Xu M, et al. Prognostic value of interleukin-6, C-reactive protein, and procalcitonin in patients with COVID-19. J Clin Virol 2020;127:104370.

5. Tan C, Huang Y, Shi F, et al. C-reactive protein correlates with computed tomographic findings and predicts severe COVID-19 early. 2020;92:856–62.

6. McFadyen JD, Zeller J, Potempa LA, Pietersz GA, Eisenhardt SU, Peter K. C-Reactive Protein and Its Structural Isoforms: An Evolutionary Conserved Marker and Central Player in Inflammatory Diseases and Beyond. Subcell Biochem 2020;94:499–520.

7. Sun Y, Dong Y, Wang L, et al. Characteristics and prognostic factors of disease severity in patients with COVID-19: The Beijing experience. J Autoimmun 2020;112:102473.

8. Yuan J, Zou R, Zeng L, et al. The correlation between viral clearance and biochemical outcomes of 94 COVID-19 infected discharged patients. Inflamm Res 2020;69:599–606.

9. Cardoso VG, Gonçalves GL, Costa-Pessoa JM, et al. Angiotensin II-induced podocyte apoptosis is mediated by endoplasmic reticulum stress/PKC-δ/p38 MAPK pathway activation and trough increased Na(+)/H(+) exchanger isoform 1 activity. BMC Nephrol 2018;19:179.

10. Ware LB, Matthay MA. The acute respiratory distress syndrome. N Engl J Med 2000;342:1334–49.

11. Gurwitz D. Angiotensin receptor blockers as tentative SARS-CoV-2 therapeutics. Drug development research 2020.

12. Pirola CJ, Sookoian S. Estimation of Renin-Angiotensin-Aldosterone-System (RAAS)-Inhibitor effect on COVID-19 outcome: A Meta-analysis. J Infect 2020;81:276–81.

13. Rothlin RP, Vetulli HM, Duarte M, Pelorosso FG. Telmisartan as tentative angiotensin receptor blocker therapeutic for COVID-19. Drug development research 2020.

14. Sproston NR, Ashworth JJ. Role of C-Reactive Protein at Sites of Inflammation and Infection. Front Immunol 2018;9:754.

15. Schulz KF, Altman DG, Moher D. CONSORT 2010 statement: updated guidelines for reporting parallel group randomised trials. BMJ 2010;340:c332.

16. Franco R, Rivas-Santisteban R, Serrano-Marín J, Rodríguez-Pérez AI, Labandeira-García JL, Navarro G. SARS-CoV-2 as a Factor to Disbalance the Renin–Angiotensin System: A Suspect in the Case of Exacerbated IL-6 Production. The Journal of Immunology 2020:ji2000642.

17. Villard O, Morquin D, Molinari N, Raingeard I, Nagot N. The Plasmatic Aldosterone and C-Reactive Protein Levels, and the Severity of Covid-19: The Dyhor-19 Study. 2020;9.

18. Wang Z, Yang B, Li Q, Wen L, Zhang R. Clinical Features of 69 Cases With Coronavirus Disease 2019 in Wuhan, China. Clin Infect Dis 2020;71:769–77.

19. Kakuta H, Sudoh K, Sasamata M, Yamagishi S. Telmisartan has the strongest binding affinity to angiotensin II type 1 receptor: comparison with other angiotensin II type 1 receptor blockers. Int J Clin Pharmacol Res 2005;25:41–6.

20. Michel MC, Foster C, Brunner HR, Liu L. A systematic comparison of the properties of clinically used angiotensin II type 1 receptor antagonists. Pharmacol Rev 2013;65:809–48.

21. Schumacher H, Mancia G. The safety profile of telmisartan as monotherapy or combined with hydrochlorothiazide: a retrospective analysis of 50 studies. Blood Press Suppl 2008;1:32–40.

22. Zhao J, Yuan Q, Wang H, et al. Antibody responses to SARS-CoV-2 in patients of novel coronavirus disease 2019. Clin Infect Dis 2020.

23. Sun B, Feng Y, Mo X, et al. Kinetics of SARS-CoV-2 specific IgM and IgG responses in COVID-19 patients. Emerging microbes & infections 2020;9:940–8.

